# Cycles of self-reported seizure likelihood correspond to yield of diagnostic epilepsy monitoring

**DOI:** 10.1101/2020.10.05.20207407

**Authors:** Philippa J. Karoly, Dominique Eden, Ewan S. Nurse, Mark J. Cook, Janelle Taylor, Sonya Dumanis, Mark P. Richardson, Benjamin H. Brinkmann, Dean R. Freestone

**Affiliations:** Graeme Clark Institute and Department of Biomedical Engineering, The University of Melbourne, Australia; Department of Medicine, St Vincent’s Hospital, The University of Melbourne, Australia; Seer Medical, Melbourne, Australia; Epilepsy Foundation, Landover, MD USA; Division of Neuroscience, Institute of Psychology Psychiatry & Neuroscience, King’s College London, London, UK; Department of Neurology, Mayo Foundation, Rochester, MN USA

## Abstract

**Objective:** Video-electroencephalography (vEEG) is an important component of epilepsy diagnosis and management. Nevertheless, inpatient vEEG monitoring fails to capture seizures in up to one third of patients during diagnostic and pre-surgical monitoring. We hypothesized that personalized seizure forecasts could be used to optimize the timing of vEEG and improve diagnostic yield.

**Methods:** We used a database of ambulatory vEEG studies to select a cohort with linked electronic seizure diaries of more than 20 reported seizures over at least 8 weeks. The total cohort included 48 participants. Diary seizure times were used to detect individuals’ multi-day seizure cycles and estimate times of high seizure risk. We then compared whether estimated seizure risk was significantly different between diagnostic and non-diagnostic vEEGs, and between vEEG with and without recorded epileptic activity.

**Results:** Estimated seizure risk was significantly higher for diagnostic vEEGs and vEEGs with epileptic activity. Across all cycle strengths, the average time in high risk during vEEG was 29.1% compared with 14% for the diagnostic/non-diagnostic groups and 32% compared to 18% for the epileptic activity/no epileptic activity groups. On average, 62.5% of the cohort showed increased time in high risk during vEEG when epileptic activity was recorded (compared to 28% of the cohort where epileptic activity was not recorded). For diagnostic vEEGs, 50% of the cohort had increased time in high risk, compared to 21.5% for non-diagnostic vEEGs.

**Significance:** This study provides a proof of principle that scheduling monitoring times based on personalized seizure risk forecasts can improve the yield of vEEG. Importantly, forecasts can be developed at low cost from mobile seizure diaries. A simple scheduling tool to improve diagnostic outcomes has the potential to reduce the significant cost and risks associated with delayed or missed diagnosis in epilepsy.

**Key Points:**

- Self-reported seizure cycles, measured from electronic diaries, correspond to outcomes of diagnostic video-EEG monitoring
- Cycle-based forecasts of seizure risk were related to both diagnostic yield and occurrence of epileptic activity during monitoring
- Personalized forecasts can be used to schedule video-EEG monitoring to coincide with periods of heighted seizure risk
- Smart scheduling of video-EEG monitoring could improve yield and reduce the burden of delayed or missed diagnoses in epilepsy

## Introduction

Video-electroencephalography (vEEG) is commonly required for the diagnosis of epilepsy syndromes^1^ and is also an important component in surgical planning. Monitoring duration for vEEG generally ranges from 1 to 14 days, although is typically less than one week^2^. Even with continuous, multi-day recording, vEEG often fails to capture seizures or relevant events, yielding an inconclusive diagnostic outcome. For instance, a study of over 100 inpatient vEEGs (with mean duration 5.6 days) showed 30% of studies failed to capture a seizure across both diagnostic and pre-surgical cases^3^, although the diagnostic yield was not directly reported. A smaller cohort study comparing inpatient and ambulatory EEG reported inconclusive diagnostic outcomes for approximately 20% of cases^4^. Other studies have found as many as 50% of inpatient EEGs may fail to record relevant events and necessitate repeat monitoring^5,6^. In addition to the significant cost of repeated hospital monitoring, delayed or incorrect diagnoses imposes a significant burden on people living with epilepsy.

Estimates of misdiagnosis rates in epilepsy vary substantially^7^, with significant heterogeneity arising from diverse clinical settings, patient cohorts, study design and clinician experience^8^. Two targeted studies from UK centers estimated misdiagnosis rates around 20%^9,10^, with psychogenic non-epileptic seizures (PNES) and syncope the most common cause of confusion^8,11^. Misdiagnosis imposes significant clinical, economic, and psychosocial costs^8,12^ and can be life-threatening for people with underlying serious cardiac arrhythmias^11^, which may be exacerbated by unnecessary AEDs. Delayed diagnosis is also costly, imposing risk of injuries and preventable accidents, including motor vehicle accidents. A large study using data from the Human Epilepsy Project found that the median time to diagnosis was over one year for people with subtle seizures, and 49 days for disruptive seizures^13^. Diagnostic delay for PNES is more severe, with a reported mean delay of around 8 years^14^. For people with PNES, this long delay places them at risk from untreated psychiatric comorbidities as well as from inappropriate medication^15^.

Given the serious consequences of delayed or missed diagnoses in epilepsy, methods to improve the yield of vEEG monitoring could be vital in reducing risks for people with epilepsy, and lead to significant cost-savings for healthcare systems. To the best of our knowledge, there are no studies that have investigated methods to systematically optimize the timing of diagnostic or pre-surgical EEG monitoring to coincide with periods when seizures (or suspected seizures) are more likely. One key reason for this is that methods to estimate future seizure likelihood have traditionally relied on EEG signals, and/or have been based on short-term (minutes to hours) prediction horizons^16^. However, it is increasingly recognized that predictable temporal patterns of seizure occurrence can be used to forecast seizure likelihood^16–18^, even days in advance^19^. Circadian rhythms have long been recognized in epilepsy and may be used clinically to schedule EEG for nocturnal or diurnal events; however, circadian rhythms have no utility for scheduling multi-day monitoring greater than 24-hours. On the other hand, longer, multi-day cycles also modulate seizure likelihood, and are well documented across human^20–23^ and animal^24,25^ studies. These multi-day rhythms have time scales over weekly, monthly and even longer seasonal or annual periodicities, and are specific to the individual^26,27^. The causes of multi-day seizure cycles remain unclear, although there appears to be no link to gender, epilepsy syndrome or seizure type^21,22,28^. We hypothesized that individuals’ seizure cycles could be used to time EEG monitoring to coincide with peak seizure likelihood.

We have shown that multi-day seizure cycles can be measured non-invasively using self-reported seizure times; and, for most people, seizure cycles measured from self-reported events correlated with their cycles of electrographic seizures^29^. There is now growing evidence that seizure diaries can be used to generate accurate, personalized forecasts of future seizure likelihood^29–31^. Diary-based forecasts are compelling due to their widespread availability, ease-of-use and low-cost implementation (typically available via free digital platforms or mobile apps)^32^. Nevertheless, concerns remain about the reliability of seizure diaries, due to underreporting, reporting non-seizure events, and other user biases^33^ and the validity and safety of forecasts developed from seizure diaries remains to be tested in a prospective setting. Nevertheless, optimizing the timing of vEEG is appealing as a low risk application for forecasts of seizure likelihood derived from seizure diaries. The aim of the study was to determine whether seizure cycles (measured from self-reported diary times) correspond to the diagnostic yield and the occurrence of epileptic activity during vEEG.

## Methods

The following sections outline a retrospective, proof-of-concept study using a database of diagnostic vEEG recordings (Seer Medical) with linked electronic seizure diaries. The study was approved by the St Vincent’s Hospital Human Research Ethics Committee (LRR 165/19). All participants provided written informed consent for their de-identified data to be used for subsequent research.

### Ambulatory video-EEG-ECG

Ambulatory video-EEG-ECG data was obtained from a large retrospective cohort of people undergoing diagnostic testing for epilepsy. For this study, we used 48 records that were also linked with seizure diaries from a freely available mobile app. No other clinical or demographic eligibility criteria were used. The range of vEEG was 1 to 10 days (mean 6.5 days). Only diagnostic reports and event labels derived from vEEG were used for this study.

Suspect epileptiform events were labelled using computer-assisted review, whereby event detection was first performed by a machine learning algorithm (see Clarke et al. 2019^34^). Events were then reviewed by expert neurophysiologists. This study only considered events that were confirmed to be either clinical seizures, or epileptiform discharges of at least 10s duration (henceforth referred to collectively as ‘epileptic activity’).

In addition to comparing vEEG with and without epileptic activity, we also compared vEEG based on diagnostic yield, using the referral indication and concluding report written by the reviewing neurologist (author MJC). Diagnostic yield was assessed by an expert neurophysiologist (author JT), who was blinded to any information about the individual’s seizure cycles or forecast.

### Seizure diaries

Self-reported seizure times were recorded in a freely available medication and seizure diary app (Seer App) that patients used before and during their vEEG monitoring and for ongoing management. For this study, only seizure times were extracted from mobile diaries. To be included in the study, users must have logged a minimum of 20 seizures (excluding seizures recorded within the same hour) and have a diary duration of at least 8 weeks.

Typically for the cohort, vEEG monitoring preceded users’ longest period of diary usage. Hence, for this study, we retrospectively extrapolated seizure cycles, and then assessed the extent that vEEG monitoring period overlapped with the high-risk period of the cycle. In this way, the study aimed to determine whether cycles of seizure likelihood were *correlated* with monitoring outcomes, rather than attempting to *forecast* monitoring outcomes. Diary seizures that were entered during the vEEG monitoring were not used to measure seizure cycles or stratify high risk periods.

### Cycle-based determination of seizure risk

We have previously shown how self-reported (diary) seizure times can be used to infer individual cycles of seizure likelihood and determine periods of low or high risk^25^. This study used the same approach. Briefly, we assessed the phase locking of self-reported seizure times to different candidate cycles using the R-value to quantify phase locking (R = 0 indicates no phase locking, while R = 1 indicates perfect periodicity).

We tested candidate cycle periods from 3 days to a maximum of 42 days, with up to two cycles selected (a ‘fast’ and ‘slow’ cycle). The fast cycle was selected from a range of 3 - 10 days and the slow cycle was selected from between 7 - 42 days. Note that circadian rhythms were not used, as vEEG covered multiple 24-hour periods. The two strongest cycles above a minimum R-value threshold were selected within each range, provided there was at least 5 days between cycles. Where the two strongest cycle periods were within 5 days of one another, the next strongest cycle was selected. The minimum R-value threshold was varied between 0.1 and 0.4 in order to explore the effect of varying cycle strengths.

For each candidate cycle period, self-reported seizure times were binned according to the phase of the cycle at which they occurred (into 20 equally sized bins from 0 to 2π). The distribution of seizure phases was then quantified using the R-value. The fast and slow cycles with the highest R-values were selected to develop a model of seizure likelihood with respect to the phase of each cycle (fast and slow).

We set a high-risk threshold for the seizure likelihood using a grid search optimization technique to maximize the number of seizures that were reported above the high-risk threshold, whilst minimizing the total time spent in high risk^23^. The cycle-based model was then extrapolated to measure the seizure likelihood within the vEEG monitoring window. This likelihood was quantified using the high-risk threshold to determine the amount of time spent in the high-risk state during vEEG.

### Validity of seizure risk estimates

The cohort was divided into distinct groups based on whether epileptic activity was recorded (‘epileptic activity’ vs ‘no epileptic activity’) and based on diagnostic yield (‘diagnostic’ vs ‘non-diagnostic’). The time spent in high risk during vEEG was compared between these groups using a one-sided t-test to determine whether the ‘epileptic activity’ and/or ‘diagnostic’ groups spent significantly more time in high risk than the ‘no epileptic activity’ or ‘non-diagnostic’ groups, respectively.

Individuals spend different overall amounts of time in high risk, so it was important to also compare the time in high risk during vEEG monitoring with each person’s baseline average time in high risk. Hence the groups were also compared based on what proportion of the cohort showed an increase in the time spent in high risk during vEEG compared to their average.

There were potential confounding factors that could contribute to differences between the diagnostic yield and/or occurrence of epileptic activity during vEEG. We assessed whether monitoring duration, self-reported seizure frequency, forecast accuracy (calculated as the percentage of diary seizures that occurred during high risk) and the baseline average time in high risk were significantly different between the diagnostic or epileptic activity groups using the same t-test statistic.

## Results

There were 48 patients included in the study (32 female) with an age range from 10 to 67 (mean 35.1) y at time of vEEG monitoring. Across the cohort, 19 out of 48 (40%) had epileptic activity recorded during monitoring and 35 out of 48 (73%) had a successful diagnostic outcome (see Table 1). There were 22 people who had vEEG consistent with focal epilepsy, 9 with generalized and 17 with a non-epileptic or undetermined vEEG. It is important to note that the distinctions between focal, generalized and non-epileptic/undetermined were based on the vEEG report and did not necessarily reflect the final diagnosis made by the treating neurologist. Additionally, the non-epileptic reports did not necessarily identify alternative conditions (i.e. PNES, syncope, sleep or cardiac abnormalities).

**Table 1.** Numbers and characteristics of people with focal, generalized, or normal/unknown vEEG. Diagnosis meant that changes observed in the vEEG were consistent with a focal or generalized epilepsy or were otherwise either normal or undetermined. ‘Mean seizure frequency’ was the cohort average of the self-reported frequency of events (per month) from users’ mobile diaries. ‘Diagnostic yield’ and ‘Epileptic activity’ show the number (proportion) of people in each cohort with diagnostic vEEG, or epileptic activity recorded during vEEG, respectively. ‘Mean forecasting accuracy’ was the cohort average of the percentage of diary seizures that were reported during estimated times of high risk.

| Diagnosis (vEEG report) | N | Mean seizure frequency (per month) | Diagnostic vEEG yield, N (%) | Epileptic activity, N (%) | Mean forecasting accuracy for diary events (%) |
| --- | --- | --- | --- | --- | --- |
| Focal | 22 | 11.1 | 19 (86%) | 7 (32%) | 75.6 (SD 12.9) |
| Generalized | 9 | 17.4 | 7 (78%) | 2 (22%) | 82.1 (SD 15.7) |
| Non-epileptic / Undetermined | 17 | 13.1 | 9 (53%) | 10 (59%) | 74.0 (SD 23.0) |

Fig 1 shows the distribution of cycle periods and strengths across people with focal, generalized and non-epileptic/undetermined EEG. The distributions of fast (3 - 10 days) and slow (7 - 42 days) multi-day cycles were similar for each group. The non-epileptic/undetermined group (n = 17) showed the strongest cycles, followed by the focal (n = 22) and generalized (n = 9) EEG groups (Fig 1A). However, the difference in the mean cycle strengths between focal, generalized and non-epileptic groups was not significant for any pairwise combination (p > 0.05 using a two-tailed t-test). The distribution of cycles was quite dispersed without clear peaks emerging at particular periods (Fig 1B). The focal group appeared to have a slight peak of people with approximately weekly cycles, while the generalized group may have a preference for monthly cycles, although the cohort was too small to draw any conclusions.

**Figure 1.**
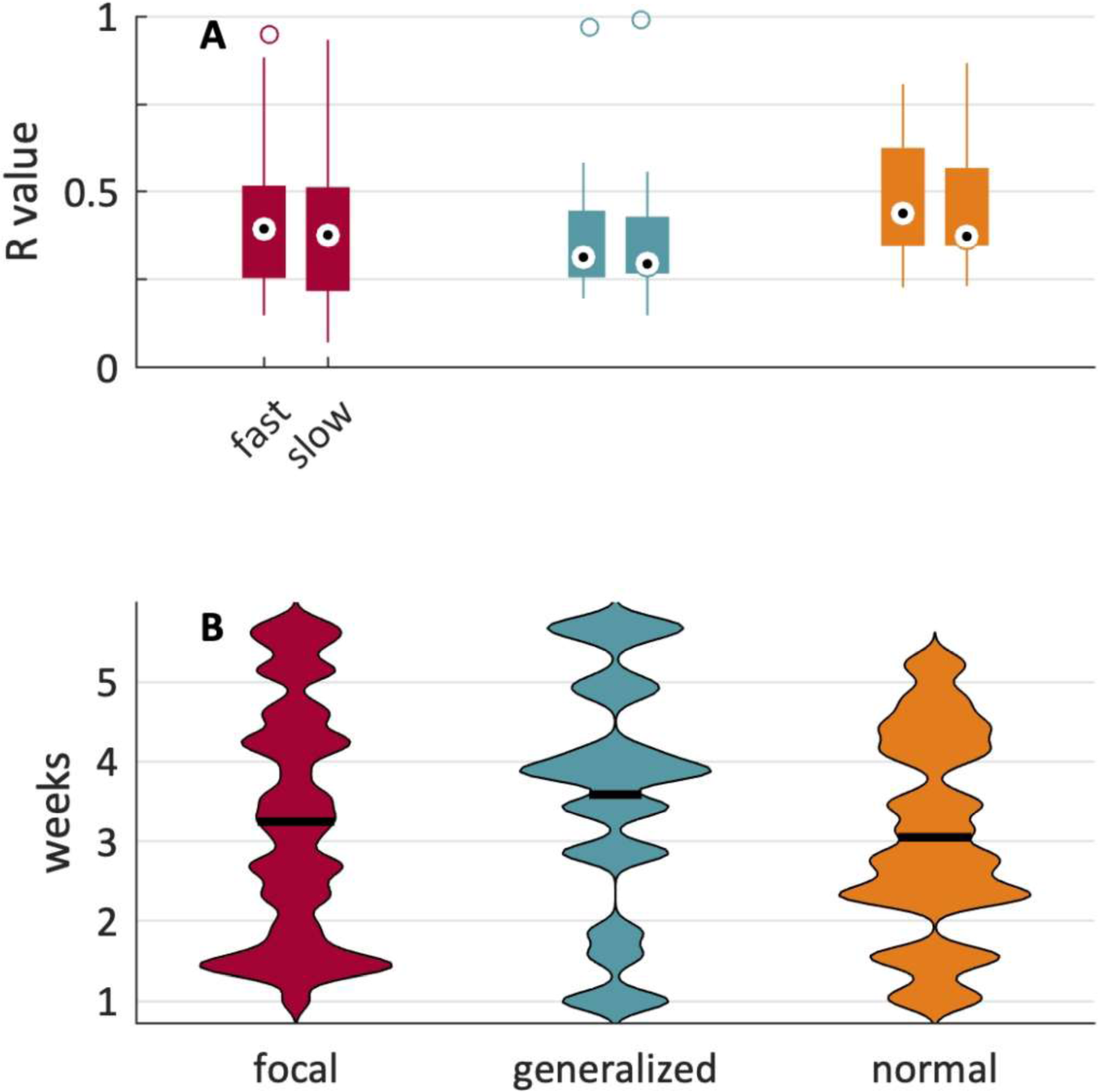
Cycle strength and distributions for focal, generalized and normal/undetermined EEG. **A.** The distributions of fast and slow multi-day cycle strengths across the cohort. Cycle strength was quantified by the R value (y-axis), where a value of 0 indicates no cycle and a value of 1 indicates perfect periodicity. Bar plots show the 25th and 75th percentiles, dots show the median, and lines show the 5th and 95th percentiles (outliers are marked by open circles). **B.** The distributions of cycle periods (y-axis) across the cohort. Violin plots represent the kernel density distributions for cycle periods. The thick black bars on each violin show the mean of the distribution. 99×99mm (300 x 300 DPI)

Fig 2 shows an example of how a seizure diary can be used to infer multi-day cycles of seizure likelihood that enable periods of high risk to be determined. The individual shown had a fast cycle of 9 days (Fig 2A) and a slow cycle of 25 days (Fig 2B). These cycles were combined to develop a continuous estimate of seizure likelihood (Fig 2C). Note that the estimated seizure likelihood fluctuates with a slower period than either cycle, due to alternating periods of constructive or destructive interference between the two cycles. In constructive interference, both cycle peaks align, leading to maximal estimated seizure likelihood, whereas destructive interference occurs when the peak of one cycle aligns with the trough of another. For this individual, the estimated seizure cycle was much stronger for their slow cycle (R = 0.52) than the fast cycle (R = 0.15, see Figs 2E and 2D). Most seizures were reported during the subject’s high-risk periods (81.3%), showing accurate performance. When the estimated cycle of seizure likelihood was projected it was found that 60.1% of vEEG monitoring time corresponded to the high-risk state, compared to their baseline time in high-risk of 35%. For this individual, focal epileptic activity was also recorded during their vEEG.

**Figure 2.**
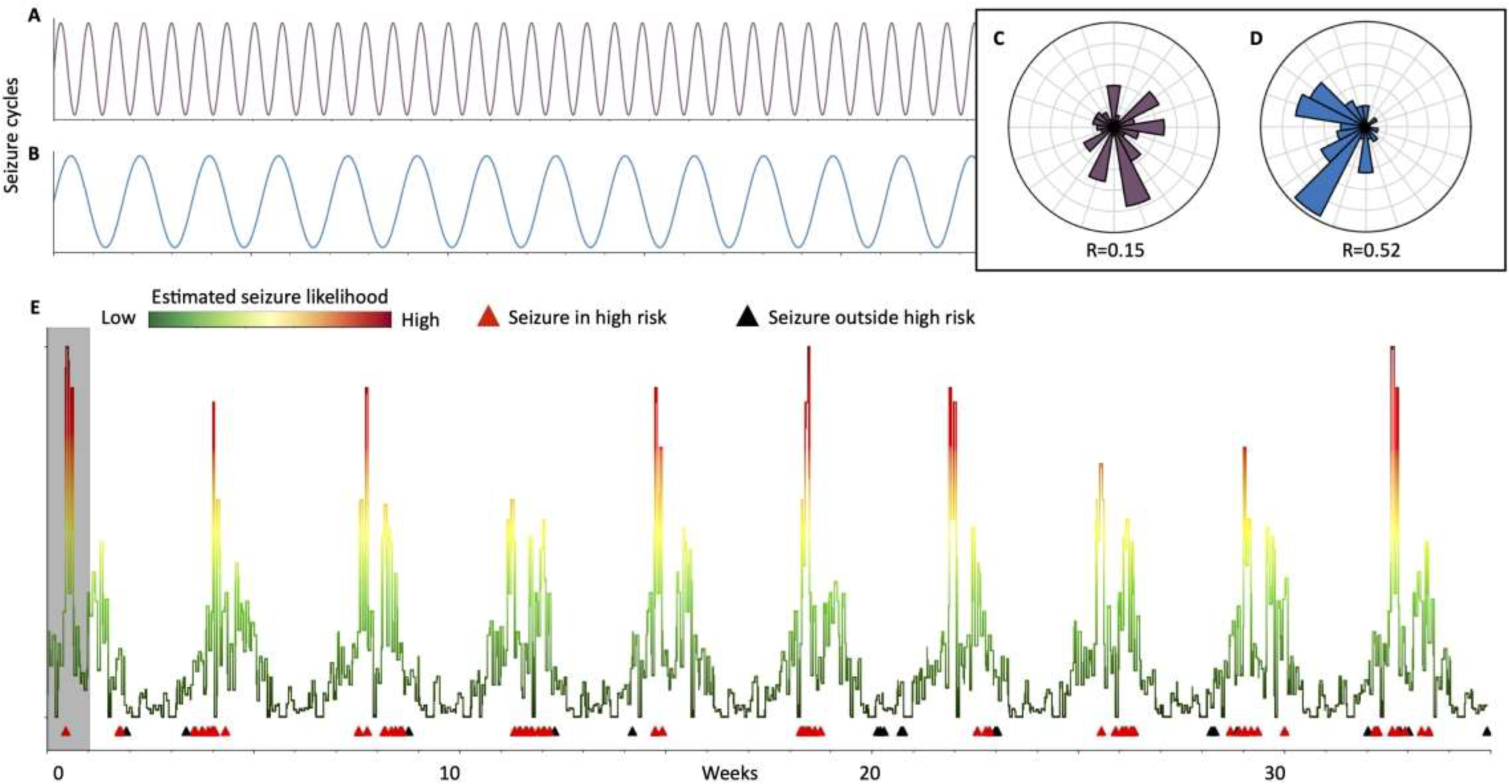
Example of an individual’s multi-day seizure cycles and correspondence with vEEG monitoring. **A,B.** Estimated fast (9 days) and slow (25 days) seizure cycles, respectively **C,D.** Circular histograms showing phase-locking of self-reported seizures to fast and slow cycles, respectively. R-values quantify the strength of phase locking. **E.** Estimated seizure likelihood. Black markers indicate diary seizures reported outside of high risk periods, and red markers indicate diary seizures in high risk periods. vEEG monitoring period shaded in grey. 255×135mm (300 x 300 DPI)

Fig 3 shows the distribution of time spent in high risk during vEEG monitoring across the cohort, at different thresholds of cycle strength (R-value). The R-value ranges from 0 to 1, with higher values indicating a stronger cycle governing the occurrence of self-reported seizure times. For each minimum cycle strength, the cohort was grouped based on whether or not epileptic activity was recorded during vEEG (Fig 3A) or based on the diagnostic yield of monitoring (Fig 3B).

**Figure 3.**
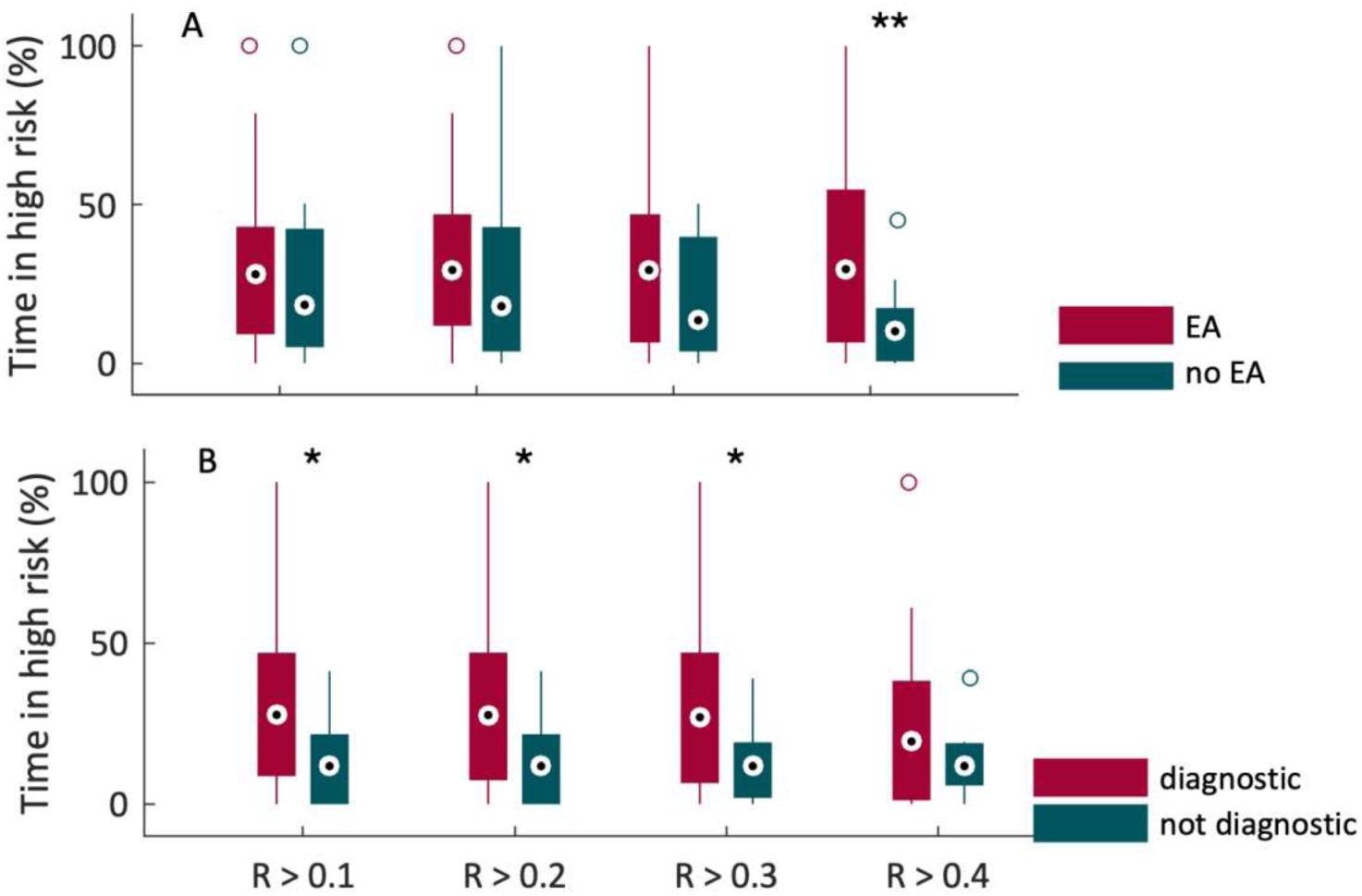
Time in high risk during vEEG across the cohort, for different cycle strengths. The distribution of the percentage of time spent in high risk during vEEG (y-axis) is compared between groups. Bar plots show the 25th and 75th percentiles, dots show the median, and lines show the 5th and 95th percentiles (outliers are marked by open circles). Group level comparisons were performed for increasing cycle strengths (minimum R-values ranging from 0.1 to 0.4). At each cycle strength, a one-sided t-test was used to compare whether the pairwise distributions showed significantly different means (** p < 0.01, * p < 0.05). **A.** Comparison of people with and without epileptic activity (EA) recorded during vEEG. **B.** Comparison of people with positive and negative vEEG diagnostic yield. 129×99mm (300 x 300 DPI)

It can be seen from Fig 3 that, for all cycle strengths, vEEG monitoring corresponded to median higher risk periods for people who had epileptic activity during monitoring as well as for people who had a successful diagnostic outcome from vEEG. In the epileptic activity versus no epileptic activity grouping, the increased time in high risk during vEEG was only significant (p < 0.05) for people with stronger seizure cycles (R > 0.4). Conversely, for the diagnostic vs not diagnostic vEEG groups, the increased time in high risk during vEEG was significant across the weaker cycles. Distribution means and p-values are given in Supplementary Tables 1 and 2.

Across all cycle strengths, the average time in high risk during vEEG was 29.1% compared with 14% for the diagnostic/non-diagnostic groups and 32% compared to 18% for the epileptic activity/no epileptic activity groups (Supplementary Table 1). This result indicates that cycles of high risk derived from seizure diaries have potential utility for scheduling vEEG in order to maximize both diagnostic yield and the likelihood of recording epileptic activity.

Fig 4 shows the percentage of people in each cohort (epileptic activity/no epileptic activity and diagnostic/non-diagnostic) where the time in high risk increased during vEEG compared to baseline. It can be seen that, across all cycle strengths, more people showed increased time in high risk during vEEGs where epileptic activity was recorded, as well as for diagnostic vEEGs. Averaged across cycle strengths, 62.5% of the cohort showed increased time in high risk during vEEG when epileptic activity was recorded (compared to 28% of the cohort where epileptic activity was not recorded). For diagnostic vEEGs, 50% of the cohort had increased time in high risk, compared to 21.5% for non-diagnostic vEEGs (Supplementary Table 2). This result provides confidence that vEEG outcomes were not simply driven by people with a higher baseline risk of seizures. Successful outcomes were more likely in people who had vEEG monitoring during periods where seizure risk was increased above baseline, highlighting the potential power of scheduling monitoring windows during these heightened risk periods.

**Figure 4.**
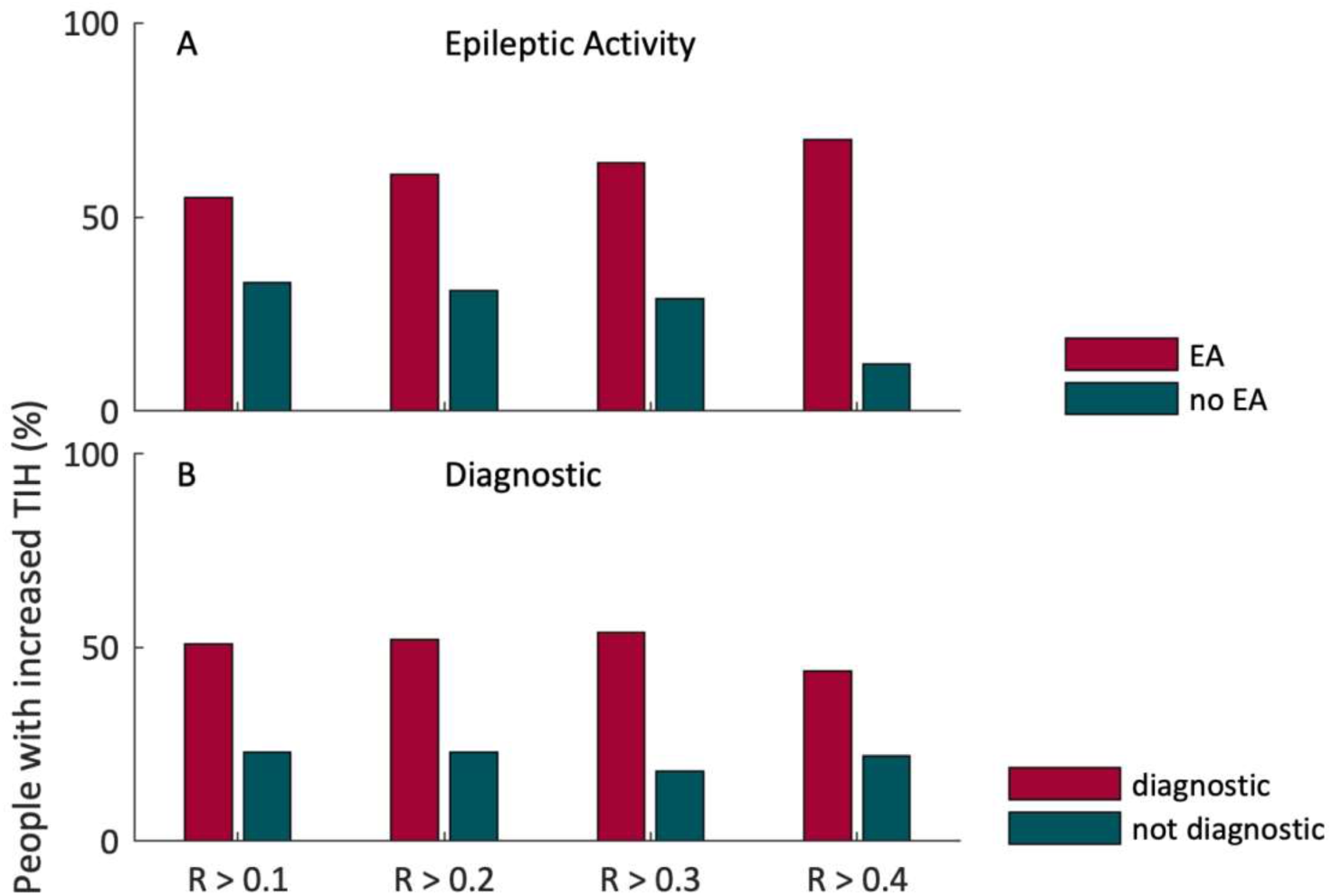
Proportion of people with increased time in high risk (TIH) during vEEG monitoring across different cycle strengths. The percentage of the cohort that showed greater time in high risk (y-axis) during vEEG compared to their baseline average time in high risk. **A.** Comparison of people with and without epileptic activity (EA) recorded during vEEG. **B.** Comparison of people with positive and negative vEEG diagnostic yield. 129×99mm (300 x 300 DPI)

In addition to the effect of baseline seizure risk, there are potential confounding factors that could affect the results of this study. We found that the duration of vEEG monitoring, seizure frequency, forecast accuracy and baseline time in high risk were not significant predictors of vEEG monitoring outcomes. Supplementary Tables 3 and 4 show these results for the epileptic activity and diagnostic yield groups, respectively.

## Discussion

This study demonstrated that the yield of diagnostic vEEG corresponded to cycles of seizure likelihood estimated from self-reported seizure times. The utility of seizure cycles was apparent regardless of whether yield was defined as recording epileptic activity, or diagnostic yield. Both the total time in high risk (Fig 3) and the proportion of people with increased risk (Fig 4) during vEEG were higher for successful monitoring outcomes. These results make a compelling case for a novel way to schedule vEEG. Currently, vEEG may be inconclusive or fail to record epileptic activity for 20-30% of cases^3,4^, often necessitating repeat monitoring. Mobile seizure diaries provide a widely available, low-cost tool that can be deployed to improve the yield of both diagnostic and pre-surgical vEEG. Given the high costs associated with in-hospital monitoring, even a small increase in yield could confer large savings. Similarly, a simple tool to improve diagnostic outcomes could reduce some of the significant risks associated with diagnostic delay or misdiagnosis in epilepsy^8^.

Cycle strength was relevant to the occurrence of epileptic activity during vEEG, with only the strongest cycles demonstrating a significant correlation between the time in high risk and whether epileptic activity was recorded (Fig 4A). This finding suggests stronger cycles of self-reported seizures were more highly correlated with the occurrence of epileptic activity during vEEG, including subclinical events. This correlation further supports our earlier work showing that cycles estimated from diary events were consistent with cycles of electrographic seizures^29^. Diagnostic yield was significantly associated with time in high risk for most cycle strengths. This is promising, since diagnostic yield is the most important outcome for epilepsy diagnosis, where less diary data may be available (leading to weaker seizure cycles).The occurrence of epileptic activity may be vital for seizure onset localization during pre-surgical monitoring either with scalp or invasive EEG, when individuals are also more likely to have an existing seizure diary. For surgical planning or management applications, the presented results suggest optimizing vEEG timing should target people with stronger, well characterized seizure cycles.

No confounding factors (monitoring duration, seizure frequency, forecast accuracy or baseline time in high risk) were found to be significantly different between the diagnostic/non-diagnostic groups at any cycle strength (Supplementary Table 4). For the epileptic activity / no epileptic activity groups (Supplementary Table 3), the forecast accuracy was significantly different (p = 0.043) between the two groups at the lowest cycle strength only (R > 0.1). Note that forecast accuracy was assessed *outside* the vEEG monitoring period (defined as the proportion of self-reported seizures that occurred during high risk states). The group with epileptic activity recorded during vEEG had less accurate forecasts (mean 70.1% of diary seizures occurred during high risk states) compared to the group without epileptic activity (mean 80.5% of diary seizures occurred during high risk states). More accurate forecasts would be expected to confer a stronger effect size in this study, since a better forecast is more likely to be well correlated with the occurrence of epileptic activity. Therefore, this result indicates that the observed effect, where estimated cycles of self-reported seizure likelihood were correlated with the occurrence of epileptic activity during vEEG, was significant *in spite of* the confounding factor and not because of it.

Forecasting accuracy and cycle strength was similar across focal, generalized and non-epileptic/ undetermined vEEG cases. The existence of cycles for non-epileptic/undetermined cases is particularly interesting as it suggests that cycles may also modulate the occurrence of non-epileptic events; although it is important to note that the current study did not differentiate between types of non-epileptic event (PNES, syncope, sleep disturbance etc.). Practically, the ability to improve diagnostic yield for PNES cases is important because of the significant diagnostic challenges for this group^14,15^. In a more general sense, the existence of multi-day cycles across other psychiatric conditions may shed light on the mechanisms of seizure cycles. Currently, causes of multi-day cycles of seizure likelihood are not understood, although candidate factors include catamenial cycles for women^35^, and possibly other hormonal factors^36,37^. Behavioral and environmental factors may also play a role, including seasonal changes and weather conditions, sleep quality, diet, exercise and stress (see ^26^ for a recent review). Psychogenic non-epileptic seizures (PNES) are linked to psychiatric comorbidities including post-traumatic stress, anxiety and depressive disorders^38^, which may show similar modulating factors as multi-day epileptic rhythms. By the same token, other episodic psychiatric conditions may adhere to slow modulation, including bipolar disorder^39,40^, depression^41,42^ and other psychopathologies^43^. Studying multi-day cycles within non-epileptic populations is likely to prove crucial to fully understand the mechanisms of epileptic rhythms.

Forecasts based on seizure cycles are an appealing option for scheduling vEEG because the repetitive nature of the cycle enables an estimate of seizure likelihood to be projected weeks or months into the future. In contrast, forecasts based on “black-box” machine learning models cannot be projected beyond the range of the available data, so are less flexible for making long-range estimates of seizure likelihood. The increasing availability of electronic diary data has recently advanced machine learning techniques for diary-based seizure forecasting^30,44,45^.

However, it is important to bear in mind that self-reported diaries provide a noisy, undersampled representation of the underlying true seizure likelihood. The likelihood model used in the current analyses, combining a fast and slow cycle, captures physiological domain knowledge that has been extensively validated to accurately describe epileptic rhythms^17,19–22^. By definition, black-box models contain no domain knowledge and simply use a large number of parameters to fit the dynamics of the training data; i.e., seizure diaries. Consequently, black-box models are more likely to fit noise or biases inherent in diaries, rather than capturing true seizure likelihood. Such biases may result in fragile forecasting models that do not generalize well to future data. Further, cycle-based models are characterized by a very small number of parameters (fast and slow cycle phase), making them less susceptible to overfitting noisy datasets with limited samples (i.e. 10s to 100s of reported events).

Due to limitations of the dataset, this study was retrospective, and assessed whether estimated cycles of seizure likelihood were correlated with vEEG outcomes, rather than performing a prospective or pseudoprospective analysis of future vEEG monitoring. However, as seizure cycles are a repeating pattern that can be projected forwards or backwards in time, retrospective analysis serves as a proof-of-concept for the prospective utility of seizure diaries to forecast optimal monitoring periods. Furthermore, individuals’ seizure cycles remain consistent over years^19,20^; so, under the assumption that estimated risk reflects these underlying cycles, estimates should be stable across time. Future work will focus on a randomized clinical trial using self-reported diary events to schedule vEEG monitoring. In line with this, it is worth noting that only around 50% of the cohort with a diagnostic vEEG and 50 - 70% of the cohort with epileptic activity actually had their monitoring during a period of increased risk compared to baseline (Fig 5). Therefore, the current results likely reflect a lower bound on the expected effect size for a prospective study where vEEG can be actively scheduled to correspond with individuals’ seizure risk. Another challenge in using seizure diaries to schedule vEEG is the minimum diary duration and/or number of seizures required. This study used an 8-week cutoff and a minimum of 20 seizures. However, in reality these limitations could be impractical, especially for people undergoing diagnostic vEEG following their first event. In practice, the use of a scheduling tool must ensure that vEEG monitoring is not delayed while waiting for diary entries.

Despite the aforementioned limitations of the study, our results show a promising avenue to develop a personalized EEG booking tool that can improve diagnostic outcomes. Such a tool represents a low-risk clinical application for seizure forecasting devices, such as the My Seizure Gauge system^25^. To follow up the present study, we aim to validate and launch a freely available seizure risk tool for people with epilepsy and medical professionals.

## Data Availability

Raw data were provided by Seer Medical (mobile app and EEG). Derived data supporting the findings of this study are available via a data sharing agreement by request to the corresponding author.

## Acknowledgements

PJK is supported by an Australian Government NHMRC Investigator Grant (EL1/1178220) MPR is supported by the MRC Centre for Neurodevelopmental Disorders (MR/N026063/1) and by the NIHR Biomedical Research Centre at the South London and Maudsley NHS Foundation Trust.

## Ethical Publication Statement

We confirm that we have read the Journal’s position on issues involved in ethical publication and affirm that this report is consistent with those guidelines.

